# Genetic liability to physical health conditions influences comorbidities in individuals with severe mental illness

**DOI:** 10.1101/2025.02.03.25320178

**Authors:** Djenifer B. Kappel, Sophie E. Smart, Michael J. Owen, Michael C. O’Donovan, Antonio F. Pardiñas, James T. R. Walters

## Abstract

**Background:** Individuals with severe mental illness (SMI), including schizophrenia and bipolar disorder, have elevated rates of physical health conditions, leading to increased morbidity and mortality. While environmental factors such as adverse effects of medication and lifestyle changes contribute to this burden, the role of genetic liability to physical health conditions remains underexplored. We assessed whether genetic risk for physical health conditions influences comorbidities in individuals with SMI and compared these effects to those observed in the general population.

**Methods:** We utilized data from two SMI cohorts from the UK: CardiffCOGS (n=721) and the National Centre for Mental Health (NCMH; n=1011). We tested whether polygenic risk scores (PRS) for six physical health conditions (high cholesterol, type 2 diabetes, hypertension, asthma, heart disease, and rheumatoid arthritis) were associated with having the corresponding condition in those with SMI. Models were adjusted for demographic and clinical covariates. Associations between psychiatric PRSs (schizophrenia, bipolar disorder, major depressive disorder, and ADHD) and presence of physical comorbidities were also evaluated.

**Results:** PRS for physical health conditions were associated with the presence of the corresponding conditions in SMI cohorts, with effect sizes comparable to those reported in the general population. Adjustments for environmental factors had minimal impact on these associations. Psychiatric PRS showed weaker and less consistent associations with physical comorbidities.

**Discussion:** This study provides robust evidence supporting the role of genetic risk in the development of common physical health conditions in individuals with SMI. Our findings indicate that the occurrence of physical health comorbidities was much more strongly associated with genetic liability to physical health conditions, than with psychiatric genetic liability. The genetic risk for physical health conditions contributes additively to environmental and clinical factors in driving comorbidities among individuals with SMI. These findings indicate there would be value in incorporating genetic risk information into predictive algorithms for physical health comorbidities in those with SMI, and that PRS should be included in research studies developing and validating such algorithms.

## Introduction

People living with severe mental illnesses (SMI), such as schizophrenia, schizoaffective disorder or bipolar disorder, often experience comorbid physical health conditions^1^. Moreover, they are likely to face poorer prognoses and increased mortality from these comorbidities compared with the general population^2,3^. The underlying causes of the elevated burden of poor physical health in SMI are not fully known. While the prevalence of physical health comorbidities is known to increase after the onset of SMI, studies have also shown that the prevalence is also elevated at the point of first SMI diagnosis^4^ implying that the comorbidities are not entirely consequences of the disorders *per se*, and thus not fully explained by treatments or secondary environmental exposures such as lifestyle changes. However, additional predisposing mechanisms, such as genetic factors, may play a role in the comorbidity risk profiles that can be seen in the SMI population. Specifically, the increased prevalence of physical health conditions in this population could be related to genetic predisposition to physical or psychiatric disorders, or even both liabilities could be jointly implicated in some way.

Research into genetic factors linked to comorbidities in those with SMI has largely focused on identifying potential pleiotropic genetic factors shared by both psychiatric and physical conditions^5-12^. Genetic correlation analyses at a population level suggest the existence of loci with pleiotropic (i.e. shared) effects on SMIs and physical comorbidities; for example genetic liability to major depressive disorder (MDD) has been reported also to predispose individuals to cardiovascular disease (CVD)^13,14^. However, the overall evidence base is limited and largely inconclusive, and it is unclear whether genetic liability to any SMI has a direct causal effect on physical health.

In contrast, studies have shown that common genetic variability associated with CVD contributes to disease risk at least as significantly as some environmental determinants, such as smoking or physical inactivity^15^. The relevance of genetic risk factors in the development of physical illnesses within the general population, may reveal broader mechanisms of disease that are equally applicable to individuals with SMI. Furthermore, studies specifically investigating the genetic liability to physical health problems in SMI rather than the general population are limited. Recent studies have primarily focused on predicting antipsychotic-related adverse effects, such as the occurrence of metabolic syndromes (e.g., elevated blood lipids^16^ or rapid weight gain^17^), rather than broader physical health morbidity. These studies suggest that individuals with higher genetically inferred risk for these traits experience more of these adverse effects after initiating antipsychotic treatment. However, these analyses have been largely limited to cohorts of individuals with schizophrenia mainly treated with antipsychotics and their conclusions cannot be straightforwardly generalized to other SMIs, psychiatric medications, or broader physical health outcomes.

In this study we aim to determine if the prevalence of a range of physical health issues in those with SMI derives from genetic liability to physical health problems, and whether those effects are similar to what has been observed in the general population. Additionally, we evaluate if these genetic effects are influenced by the concomitant presence of known risk factors for physical illnesses.

## Methods

### Samples

We analysed genetic and phenotypic data from two cohorts ascertained by Cardiff University. The first, CardiffCOGS^18,19^, is a clinically ascertained sample of individuals with a diagnosis of SMI, recruited from out-patient, in-patient and voluntary sector mental health services in the UK between 2007 and 2017. Participants were interviewed using the Schedules for Clinical Assessment in Neuropsychiatry (SCAN)^20^, lifetime psychiatric diagnoses were assigned using DSM-IV^21^ and/or ICD10 ^22^criteria, and additional data derived from clinical records, participants were also invited to provide a blood sample for genetic analysis. In this study, we included participants with schizophrenia, schizoaffective disorder depressed type, delusional disorder, schizophreniform disorder – the schizophrenia spectrum group - or bipolar disorder type 1 or 2 and schizoaffective disorder bipolar type - the bipolar spectrum group – in our SMI cohort.

The second cohort, the National Centre for Mental Health (NCMH) ^23^, was recruited from health care services, voluntary organisations or via public advertisement from 2012 to 2021 ^23^. A standardised assessment was administered by trained researchers; participants were asked whether a doctor or mental health professional had ever diagnosed them with a mental health diagnosis from a list of potential psychiatric diagnoses ^24^. Additionally, a subset of participants (N=105) participated in a research interview based on the SCAN and their SMI status was assigned using DSM-IV and/or ICD-10 criteria. In this study, we used the subsample of the NCMH cohort who completed the standardised assessment, had a SMI diagnosis, and who provided a blood sample for genetic analyses. Individuals reporting that their primary psychiatric diagnosis as given by a clinical team was schizophrenia, schizoaffective disorder depressed type, delusional disorder, schizophreniform disorder were included in the schizophrenia spectrum group, and those with a bipolar disorder type 1 or 2 and schizoaffective disorder bipolar type diagnosis were part of the bipolar spectrum group. A previous study using this cohort reported good validity of the self-reported SMI diagnoses when assessed against a complete research interview using the SCAN instrument ^24^.

NCMH was approved by the Health Research Authority and Wales Research Ethics Committee (REC) (REC reference: 16/WA/0323), and CardiffCOGS by the NHS REC (REC reference: 07/WSE03/110). All participants provided written informed consent.

### Physical Health Comorbidities

For both cohorts, additional information on basic demographics and lifetime clinical and lifestyle factors (i.e., sex, age, smoking history, and use of the antipsychotic clozapine) were extracted from the interviews.

Self-reported information on physical health conditions was collected consistently across both cohorts. Participants were asked to report if they had ever been told by a health care professional that they had one or more of 20 common physical health problems. The reliability of this self-reported measure of physical health conditions has been previously validated by our group against electronic health records^9^.

From the complete list of available conditions, we analysed 6 physical health conditions known to have an increased prevalence in individuals with SMI^4,9,25^ - high cholesterol, type 2 diabetes, hypertension, heart disease, asthma, and rheumatoid arthritis (Supplementary Table 2). These conditions were selected due to their consistent, specific, and comparable assessment across both cohorts, and sufficient frequency (minimum 30 cases in each cohort). We removed participants for whom most physical health data outcomes were missing (more than 4 out of 6 outcomes).

### Genetic Data

Participants from both cohorts provided blood or saliva for DNA extraction. Genomic data were obtained using the Illumina OmniExpress genotyping platform for CardiffCOGS and either the Illumina PsychArray or Illumina GSA platforms for NCMH. Quality control (QC) and imputation were performed uniformly using the DRAGON-Data QC pipeline (v.2.0)^26^ and the Haplotype Reference consortium^27^ respectively.

To identify related individuals within and between samples and to derive genetic principal components (PC), we used a subset of linkage disequilibrium-pruned common SNPs with high imputation quality (MAF > 0.05, INFO > 0.9, 500kb window, pairwise r2 < 0.2). After relatedness analyses using the DRAGON-Data QC pipeline, one individual from each pair assumed to be duplicates (kinship coefficient > 0.98) or related (kinship coefficient > 0.1875; equivalent to second degree relative) was removed at random. If duplicates or relatives were found between CardiffCOGS and NCMH, the participant in CardiffCOGS was retained. Genetic ancestry was assessed using ancestry informative markers in a linear discriminant analysis framework^28^ integrated in the DRAGON-data QC pipeline^26^.

After quality control, complete phenotypic and genetic data were available for 721 CardiffCOGS and 1011 NCMH participants.

### Polygenic Risk Scores

Using the largest available genome-wide association study (GWAS) summary statistics, we calculated 6 polygenic risk scores (PRS) for physical health traits and 4 PRS for psychiatric disorders: coronary artery disease (CAD)^29^, type 2 diabetes^30^, asthma^31^, rheumatoid arthritis (RA)^32^, systolic blood pressure (SBP)^33^, LDL-cholesterol (LDL-C)^34^, attention-deficit/hyperactivity disorder (ADHD)^35^, bipolar disorder (BD)^36^, major depressive disorder (MDD)^37^ and schizophrenia (SCZ)^38^. Since CardiffCOGS contributed to the schizophrenia GWAS, we use a custom schizophrenia GWAS generated by the Psychiatric Genomics Consortium which excluded this sample. The NCMH cohort was not included in any of these GWAS studies. The full list of GWAS summary statistics used is detailed in Supplementary Table 1.

All GWAS summary statistics were standardised and quality-controlled with the MungeSumstats^39^ package. This process included LiftOver to GRCh37 coordinates when necessary, mapping to dbSNP build 155, and filtering for multi-allelic, allele-mismatched, strand-ambiguous SNPs, indels, SNPs with missing test statistics, and variants not present in the 1000 Genomes version 3.

Following recommendations from the Global Biobank Meta-Analysis Initiative (GBMI) ^40^ for summary statistics of predominantly European ancestry, we used the PRS-CS software ^41^ with the European subset of the 1000 genomes^42^ as a LD reference to compute PRS. The PRS-CS algorithm was used with default parameters though the Markov Chain Monte Carlo procedure was run for 25000 iterations with 10000 iterations of burn-in, and automatically inferring the shrinkage parameter (--phi=auto). The PRS-cs recalculated summary statistics were then used to derive individual level PRS for all psychiatric and physical conditions with the scoring function in PLINK 2.0. In this step, SNPs with a minor allele frequency of <10%, Hardy-Weinberg equilibrium (p <1×10^−6^) or variant call rate (<98%) were removed from the CardiffCOGS and NCMH genotypic data prior to PRS scoring. Additionally, the extended major histocompatibility complex region (25–34Mb) was excluded from most PRS calculations, due to its complex LD structure, except for rheumatoid arthritis because of the region’s significant impact on variance explained by common genetic variants in this particular phenotype ^43^. PRS were scaled to a mean of zero and standard deviation (SD) of one for subsequent association analyses.

### Statistical Analysis

We conducted a within-case genetic association study to assess the relationship between PRS for psychiatric and physical health conditions and the development of physical comorbidities in two independent SMI samples. We hypothesized that each physical health PRS would be associated with its corresponding health problem and explored whether psychiatric PRS were linked to any physical health outcomes (Figure 1).

**Figure 1:**
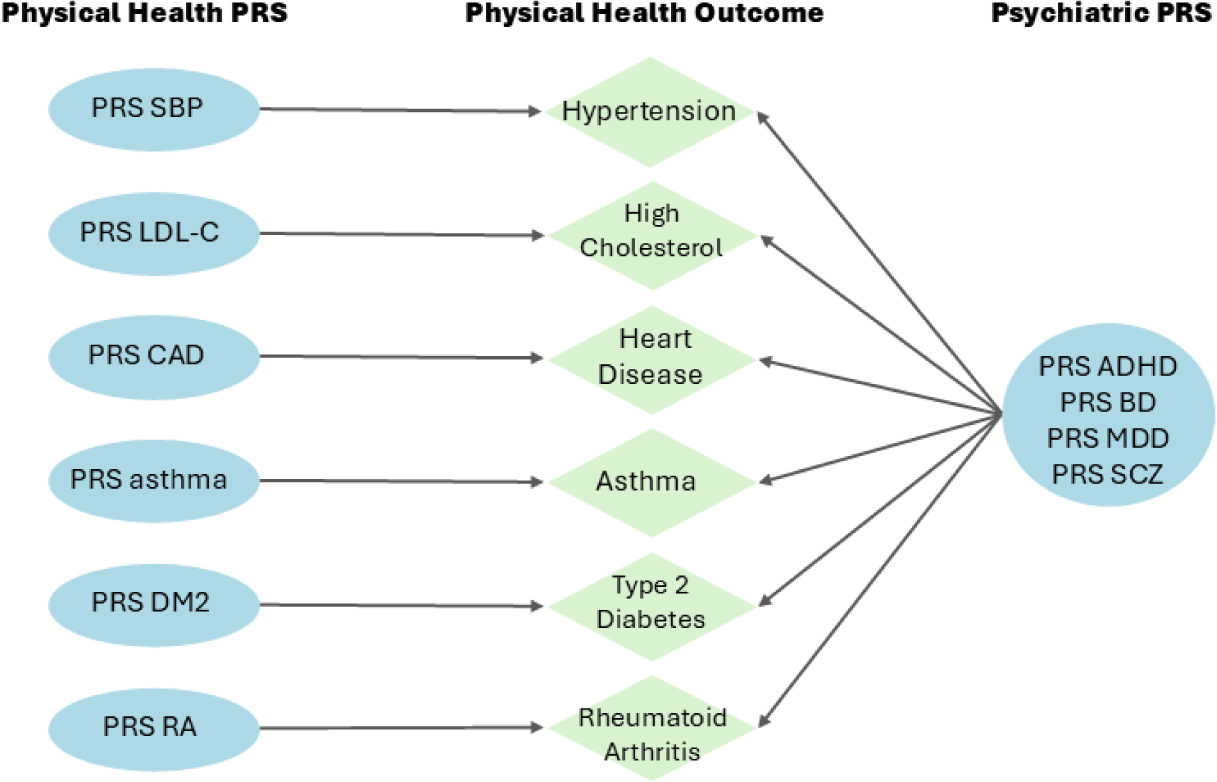
Study design diagram. The diagram illustrates the analyses performed between polygenic risk scores (PRS) and physical health outcomes in severe mental illness. Arrows represent associations tested between PRS and health outcomes. The left section shows PRS for physical health traits - systolic blood pressure (SBP), low-density lipoprotein cholesterol (LDL-C), coronary artery disease (CAD), asthma, type 2 diabetes (DM2), and rheumatoid arthritis (RA) - and their corresponding physical health outcomes in the middle section -Hypertension, High Cholesterol, Heart Disease, Asthma, Type 2 Diabetes and Rheumatoid Arthritis. The right section shows PRS for four different psychiatric conditions - attention-deficit/hyperactivity disorder (ADHD), bipolar disorder (BD), major depressive disorder (MDD), and schizophrenia (SCZ) – with analyses being performed with each PRS association with each of the individual’s physical health outcomes.

Logistic regression analyses were performed using the glmrob function from *robustbase^44^* in R version 4.4, allowing for robust estimation of standard errors given the low prevalence of certain physical health outcomes in these samples. Given our study design included individuals with mixed diagnosis SMI, we assessed if any of the physical health conditions analysed were more prevalent in the schizophrenia or bipolar spectrum samples. As we did not find significant associations between diagnosis and physical health outcomes that replicated across the CardiffCOGS and NCMH samples, we proceeded with a joint analysis of SMI (Supplementary Table 2). However, in the NCMH sample, differences in the proportions of physical health conditions between those with schizophrenia spectrum and bipolar spectrum diagnoses were observed for a few outcomes (Supplementary Table 2), which prompted the inclusion of diagnosis as a covariate in sensitivity analyses described below.

Primary baseline models were controlled for sex, age at assessment and the top 10 genomic principal components to account for population stratification in each cohort. Results are reported as effect estimates and their 95% confidence intervals per SD increase in the PRS. Nagelkerke’s pseudo-R2 was calculated to estimate the proportion of phenotypic variance explained by each of the PRSs. As this method is not implemented for the *glmrob* models described above, generalised linear models with traditional standard errors were fitted for this procedure. We also used a test for interaction (or moderators)^45^ to examine the concordance of the effect sizes (regression slopes) from each cohort. This is also called a test for moderators in the statistical literature.

For secondary fully adjusted models, we included additional covariates relevant to the SMI population. These included the diagnosis spectrum (schizophrenia or bipolar), a history of smoking or a history clozapine use, as this antipsychotic is enriched in the CardiffCOGS sample ^18^ and is associated with multiple comorbidities including weight gain and metabolic syndrome ^46^. This approach enabled us to evaluate the independent effects of higher comorbidity genetic risk from those more likely related to these environmental exposures. For models with significant associations for both PRS and environmental covariates, we performed follow-up analyses to test for interaction effects between PRS and environmental exposure.

Lastly, to examine the overall impact of the combined genetic liabilities from physical and psychiatric disorders on phenotype variability in our SMI samples, we extracted the residuals from the physical health PRS fully adjusted models described above rank-normalised those residuals and then analysed if including genetic liabilities to psychiatric disorders improved any unexplained phenotype variability using linear residual regression models including all the covariates in this model. This approach was preferred to the inclusion of additional PRS covariates in the fully adjusted models as, given the low prevalence of some of our outcomes and the resulting small sample size of their case subsets, it can avoid failures in the model fitting process caused by sparse data (“separation”)^47^.

The Benjamini-Hochberg false discovery rate (FDR) method was used to correct for multiple testing at a P^FDR^ ≤0.1 threshold^48^ within each independent cohort. Where relatively low frequency of a comorbidity resulted in computational issues (e.g. singular model fits and separation), we determined whether single covariates were responsible for the problems, and adapted the regression models to each particular case^47^. As examples of this, adjusted models for heart disease in CardiffCOGS do not include the covariate smoking, and due to the population stratification of rheumatoid arthritis in NCMH (all affected individuals are on a restricted area of the PC-space – Supplementary Figure 1), those specific analyses were restricted to individuals of European ancestry-only).

### General Population Comparison

To examine whether the results for PRS association with physical health problems in individuals with SMI were comparable with results from the general population, we extracted the association statistics from studies analysing the impact of physical health problem PRS on physical health condition susceptibility in biobanks and the general population. Specific details about the original studies regarding polygenic score development and phenotypes analysed are given in Supplementary Table 3. When multiple compatible studies were found, or a study analysed more than one sample, we conducted a meta-analysis of those results using a fixed effect model weighted by standard error, with the package *metafor^49^* in R. Comparisons of PRS effect sizes from the results in CardiffCOGS and NCMH with those extracted from studies in the general population were made by evaluating the overlap or lack thereof of 84% confidence intervals, equivalent to a two-tailed two population test with alpha = 0.05 ^50^.

Supplementary Table 3 provides detailed estimates of effect sizes retrieved from the literature, alongside the results of the meta-analysis and a direct comparison with the findings from our two cohorts. For certain traits, multiple analyses from the same biobank were included in the meta-analysis due to methodological differences across studies. These duplicates were retained to provide a comprehensive representation of the variability in the literature. As a sensitivity analysis, we also conducted a meta-analysis in which only one analysis per biobank was included but found no significant differences in the effect sizes or the overall interpretation of the results.

## Results

Participants from CardiffCOGS (N=721) had a mean age at interview of 43.65 (SD=12.05) and 267 (37.03%) were female. The majority - 639 (88.63 %) - had a schizophrenia spectrum diagnosis (schizophrenia, schizoaffective disorder depressed type, delusional disorder, schizophreniform disorder), while 82 (11.37 %) participants had a bipolar disorder (type 1 or 2) or schizoaffective disorder bipolar type diagnosis. NCMH participants included in the analyses (N=1011) had a mean age of 47.6 (SD=13.69) years at the time of interview, and 547 (54.10 %) were female. In this cohort, 388 (38.38 %) had a schizophrenia spectrum diagnosis, and 623 (61.62%) had a bipolar spectrum diagnosis. Supplementary Table 2 presents the frequencies of each physical health condition by cohort and diagnostic group.

### Physical health PRS and physical comorbidity in SMI

All six physical health PRS analysed were significantly associated with the presence of their corresponding physical comorbidity in at least one or in both SMI cohorts (Figure 2 and Supplementary Table 4). Significant PRSs explained between 1.4%–6.5% of the observed variability of each phenotype (Supplementary Table 5).

**Figure 2:**
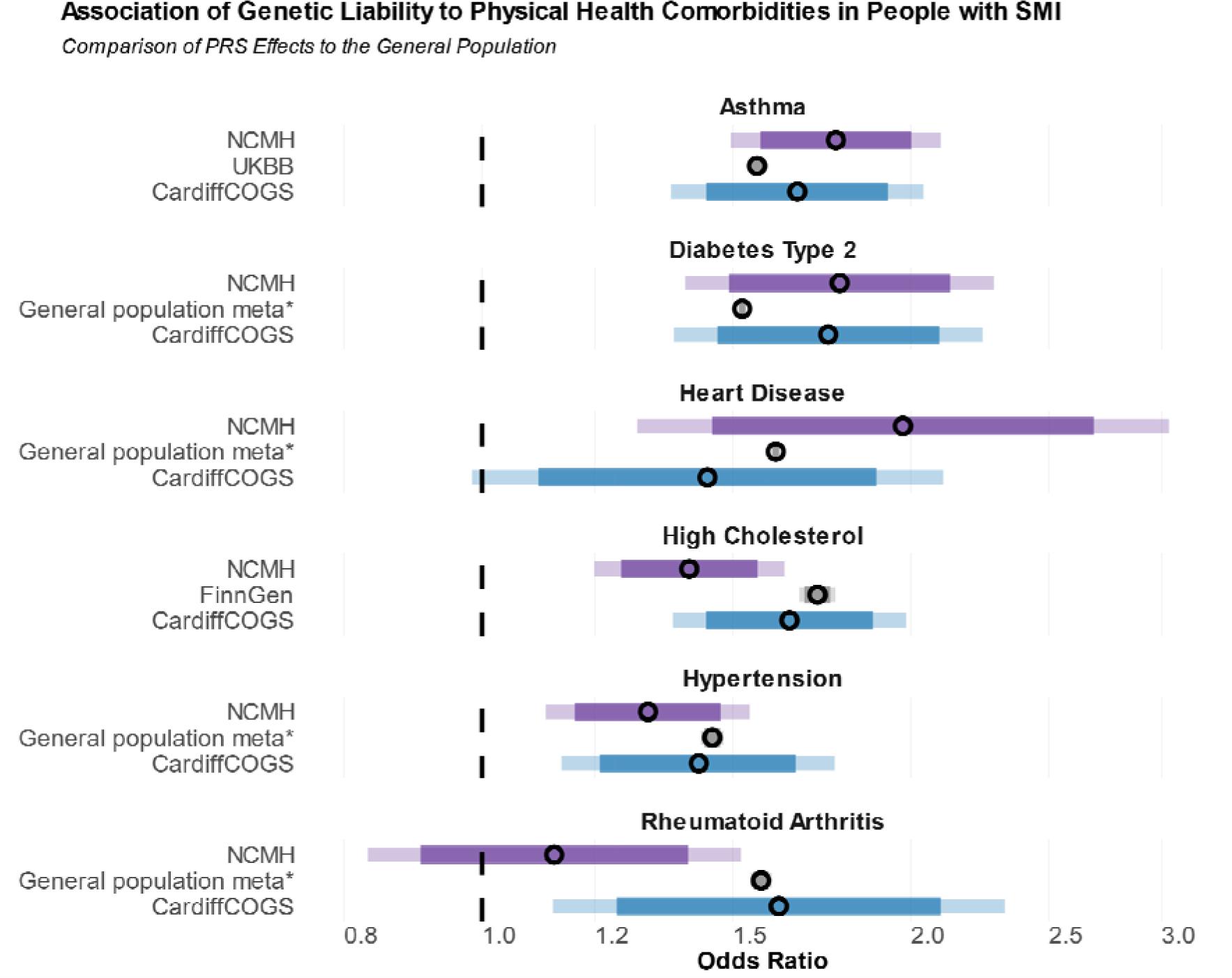
Associations between genetic liability (PRS) to physical health comorbidities and their occurrence in two cohorts of individuals with severe mental illness (NCMH N=1011; CardiffCOGS N=721), and comparison of effects observed in the general population. Results are presented as odds ratios and 84% confidence interval (darker) and 95% confidence interval (lighter). Effect sizes are derived from logistic regression models adjusting for sex, age, and 10 first principal components, details in Supplementary Table 3 and 4.

We further evaluated if the effect of the PRSs were attenuated in models adjusting for well-established clinical and demographic environmental risk factors of physical conditions (sex, age, smoking, and clozapine use – Supplementary Table 4). In these adjusted models, we observed minimal changes to the logistic regression estimates and the associations between the PRS and their respective comorbidities remained statistically significant (FDR < 0.1) in at least one cohort, except for the association of PRS-CAD with heart disease (Figure 3 and Supplementary Table 4). Heart disease had the lowest prevalence among the comorbidities analysed, with 35 and 42 individuals reporting heart problems in CardiffCOGS and NCMH, respectively. The limited sample size in this group may have reduced statistical power, particularly when adjusting for covariates.

**Figure 3:**
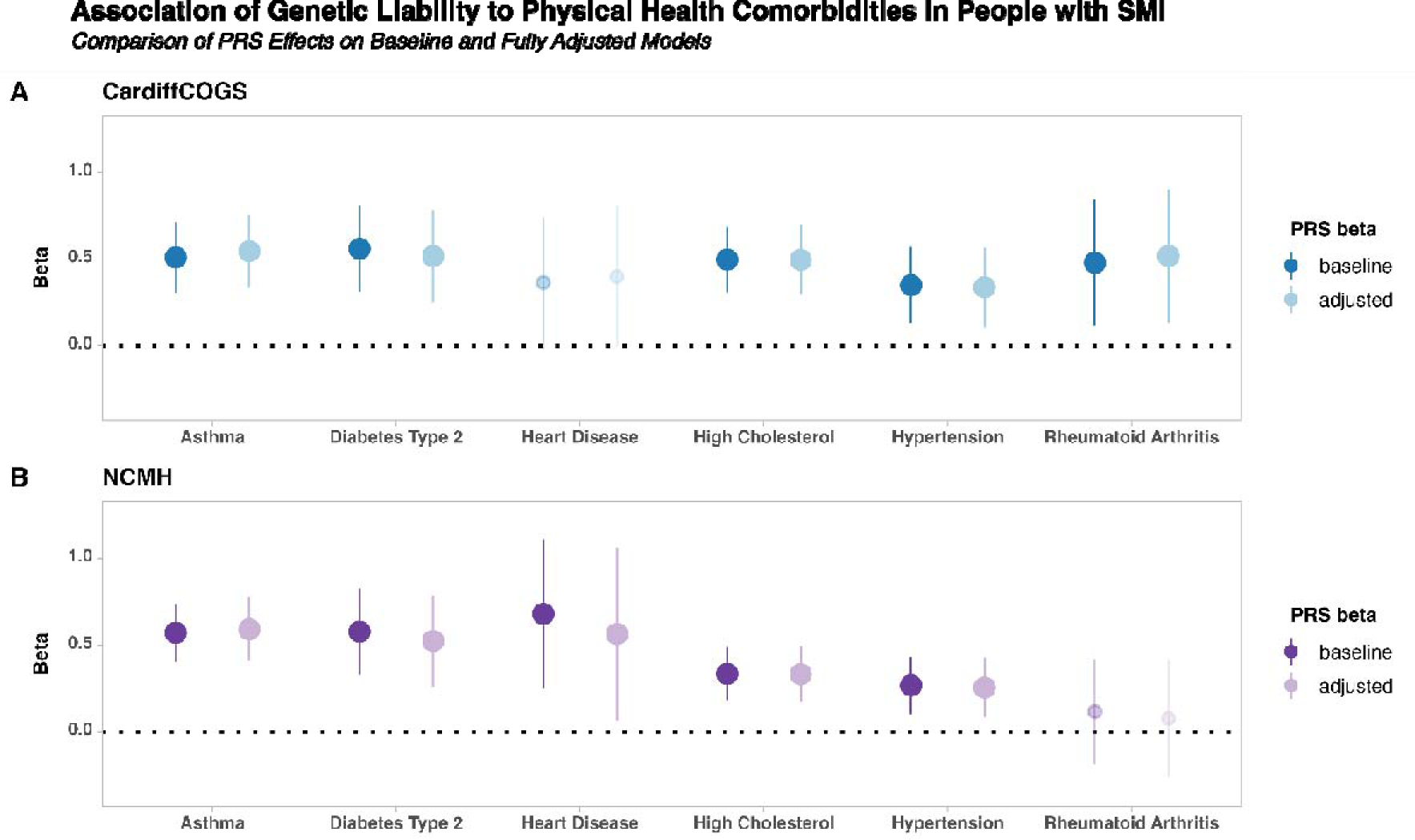
Associations between genetic liability to physical health (PRS) comorbidities (independent variable) and the corresponding comorbidity in two models adjusting for additional environmental covariates. The effect estimates are provided as beta coefficients with 95% confidence intervals. The results are presented for the baseline models adjusted for sex, age, and 10 first principal components, and the fully adjusted models with additional covariates smoking history, SCZ or BD spectrum diagnosis, and use of the antipsychotic clozapine. Larger dots and no transparency indicate that the results are statistically significant at P < 0.05, and FDR <0.1 in at least one model in each SMI cohort. Details are provided in Supplementary Table 4.

Of the identified non-genetic risk factors, age and sex showed consistent associations across physical health outcomes and clozapine was associated with higher LDL cholesterol and Type 2 diabetes (Supplementary Table 4). However, we did not detect an interaction effect of physical comorbidities PRS and clozapine, as it would have been expected under a ^51^_￼_^50^_￼_ if the effects of clozapine on its relevant comorbidities were large enough to mask any risk or protective effects conferred by the PRS (Supplementary Table 6).

### Psychiatric PRS and physical health comorbidity in SMI

We next tested whether genetic liability to four psychiatric disorders (ADHD, BD, MDD and SCZ) was associated with physical health comorbidity in people with SMI. While we identified some associations (FDR < 0.1) (Figure 4, Supplementary Table 7), the overall evidence for an effect of psychiatric PRS on comorbidity outcomes was limited and inconsistent. We observed significant differences in effect estimates between CardiffCOGS and NCMH for the associations of PRS-SCZ and both high cholesterol and rheumatoid arthritis, suggesting heterogeneity in these associations across cohorts.

**Figure 4:**
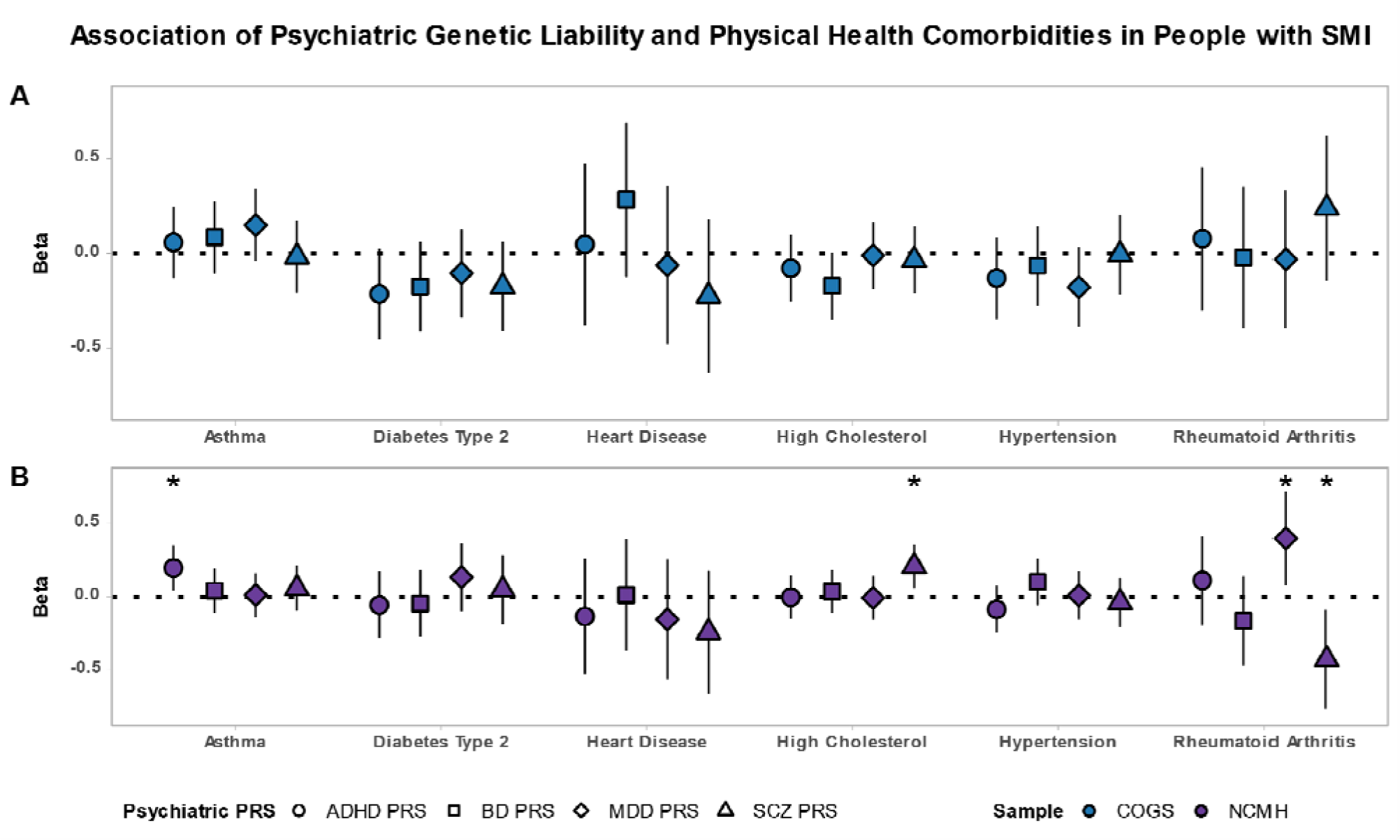
Associations between physical health comorbidities and four psychiatric PRS in two cohorts of severe mental illness (NCMH N=1011; CardiffCOGS N=721), after correction for sex, age, and 10 principal components. The symbols indicate the estimated regression effect size for each of the analysed PRS and comorbidity outcomes and lines indicate 95% confidence intervals. Asterisks indicate where a significant association (at FDR < 0.1) was observed, additional details in Supplementary Tables 7 and 8.

Given the strong association between physical health trait and their corresponding comorbidities identified above, we also analysed if the psychiatric PRS could help explain additional phenotypic variability in the occurrence of physical problems in those with SMI (Supplementary Table 8). Our results suggest that the PRS-SCZ might contribute to the risk for high cholesterol, acting additively to other genomic and environmental factors; however, the significant heterogeneity observed between our SMI cohorts warrants further scrutiny to understand if these are not related to factors specifically related to the population analysed.

## Discussion

Many of the most common causes of morbidity and mortality in the general population, such as type 2 diabetes, dyslipidaemia and cardiovascular disease, among others, are more highly prevalent in individuals with SMI^1,4,52^. The elevated rates of these comorbidities in the SMI population are often linked to demographic and lifestyle characteristics in those with SMI, such as poor diet, sedentary behaviour, smoking, and to the side effects of pharmacological treatment with antipsychotics, mood stabilisers, and antidepressants^53^. Here, we evaluated if, in addition to the known environmental factors associated with the development of comorbidities in SMI, biological mechanisms such as genetic risk associated with psychiatric and physical illness can play a role in the development of physical health problems in those with SMI. In this study we observed that in people with SMI, the occurrence of high cholesterol, type 2 diabetes, hypertension, asthma, heart disease and rheumatoid arthritis were statistically significantly associated with genetic liability to these conditions.

Given previous results suggesting that phenotypic correlations between SMI and physical comorbidities seems to be largely driven by environmental factors^14^, we also explored if the effects of PRS of physical health problems and the occurrence of these outcomes in our samples could be modified by environmental factors known to increase the risk for comorbidity in those with SMI. Our inclusion of covariates, such as smoking and medication use, did not significantly alter the observed associations between PRS for physical health conditions and the corresponding clinical outcomes. Moreover, by comparing the effect size estimates for physical health PRS and comorbidities obtained in our two SMI cohorts to estimates from studies performed in biobanks, we found that genetic variants associated with physical disorders confer a similar risk of comorbidities in individuals with SMI and in the general population. These findings suggests that the genetic component of physical health risk operates through mechanisms that are partially independent to the causes and clinical context of SMI.

While some prior studies have proposed a link between psychiatric genetic liability — particularly for MDD — and cardiovascular disease^12-14^, our findings did not reveal consistent evidence of such associations in people with SMI. The effects of psychiatric PRS on comorbid physical conditions were weaker and more variable between our cohorts, and in some cases did not offer additional contribution to the explained variance of the comorbid physical condition. This suggests that direct links between psychiatric genetic risk and physical health may not be as strong or consistent as previously reported ^5,12,13^, and these relationships may be influenced by other unmeasured variables or population-specific factors. The absence of a robust association between psychiatric genetic risk and physical health outcomes suggests that direct effects of psychiatric genetic liability may have a limited impact, however, we cannot rule out that indirect effects of the SMI — such as the impact of medication, disorder severity, and lifestyle factors — affect the development of comorbidities.

Recognizing the high prevalence of comorbid physical health conditions in individuals with SMI, there is a drive for services to integrate physical health monitoring, including metabolic and cardiovascular risk assessments, into their clinical management plans for people with SMI ^1,54,55^. Although risk assessment tools for physical health conditions are widely used in the general population, and there is increasing support for the inclusion of genetic risk factors in those ^15,56-58^, it is unclear whether these tools are equally applicable to individuals with SMI^59,60^. Recent longitudinal studies have suggested that individuals with SMI carrying higher genetic risk for physical and/or metabolic conditions were more likely to experience cholesterol level and BMI increases during pharmacological treatment with antipsychotics^16,17^, with genetic liability to these traits accounting for a substantial portion of this variance. Our findings contribute to the evidence that genetic liability, pharmacological treatment, and environmental factors independently influence the development of physical health problems in this specific population^16,17^. The validation and integration of PRSs into clinical practice could enable more personalised interventions, where genetic risk scores serve as complementary tools for early screening and treatment optimisation in individuals predisposed to physical health problems ^61,62^. Our findings provide initial evidence that strategies to address comorbid health problems in individuals with SMI at particular risk of those comorbidities should account for genetic factors for those comorbidities rather than focus only on well-known environmental risk factors, such as antipsychotic use, illness severity, and lifestyle choices. Importantly, we also show that the effects of genetic liability to physical health problems appears to be no different from what is observed in the general population. This suggests that, if genetic information is integrated into stratification and prediction algorithms in the general population, it could be equally valuable for individuals with SMI, particularly those at elevated risk from exposure to known environmental risk factors such as medication use and smoking.

## Limitations

While our study provides robust evidence for the role of genetic risk in physical comorbidities among individuals with SMI, several limitations should be acknowledged. First, our design does not allow us to examine if individuals with SMI present significantly higher genetic liability to comorbidities than a similar sample without psychiatric disorders, and if this would explain part of the elevated prevalence of some physical health problems in the SMI population.

Second, despite our efforts to control for confounding factors, we were unable to account for many important variables, such as BMI, weight changes, and socioeconomic status, which are known to affect the occurrence of physical comorbidities.

Third, while we tried to avoid excluding participants based on ancestry, the majority of our sample is comprised of individuals of European descent and at least one of our analyses had to be restricted to this sub-population. We used multi-ancestry GWAS summary statistics where available, but the generalizability of our findings to non-European populations remains uncertain. Future studies should replicate these findings in more diverse cohorts to ensure broader applicability.

Finally, this study focuses on a specific subset of physical health conditions—namely, high cholesterol, type 2 diabetes, hypertension, asthma, heart disease, and rheumatoid arthritis. While these six categories are prevalent and clinically relevant, other physical health conditions not included in our analysis could impact individuals with SMI differently, potentially leading to alternative insights or highlighting additional areas where genetic or environmental factors influence physical health comorbidities in those with SMI. Further research is needed to explore a wider range of physical health conditions in SMI populations, particularly those that are understudied yet may hold significance for this population’s overall health and quality of life.

## Conclusion

This study provides strong evidence supporting the role of genetic risk in the development of common physical health conditions in individuals with SMI. Our findings indicate that the occurrence of these comorbidities is significantly more impacted by the genetic liability to physical health conditions than of psychiatric genetic liability. These genetic effects appear to act independently to established environmental risk factors in contributing to comorbidity development. Further research is required to determine whether these findings can inform stratification and prediction models, with the potential to enhance the development of personalized medicine approaches in psychiatric care.

## Data Availability

All data produced in the present study are available upon reasonable request to the authors

## Acknowledgments

We thank the participants, clinicians, lab staff and field team for their invaluable contributions to the NCMH and CardiffCOGS studies. The National Centre for Mental Health (NCMH) is a collaboration between Cardiff, Swansea and Bangor Universities and is funded by Welsh Government through Health and Care Research Wales. We thank Alex Evans, Lesley Bates, Catherine Millson and Lucinda Hopkins (Cardiff University) for laboratory sample management, Daniel Oakes, Sarah Knott and Ian Jones (NCMH) for NCMH data management and preparation, and the Schizophrenia Working group of the Psychiatric Genomics Consortium for providing leave-Cardiff samples-out GWAS summary statistics. This study was supported by the European Union Horizon 2020 program (REALMENT; Grant No. 964874). The overall research was also supported by grants from the Medical Research Council to Cardiff University: Program (MR/P005748/1, MR/Y004094/1), Project (MR/L011794/1, MC_PC_17212) and the Brain and Genomics Hub of the Mental Health Platform (MR/Z503745/1). We acknowledge the support of the Supercomputing Wales project, partly funded by the European Regional Development Fund (ERDF) via the Welsh Government. The funders had no further role in study design; in the collection, analysis and interpretation of data; in the writing of the report; and in the decision to submit the paper for publication.

---

**Supplementary Figure 1:**
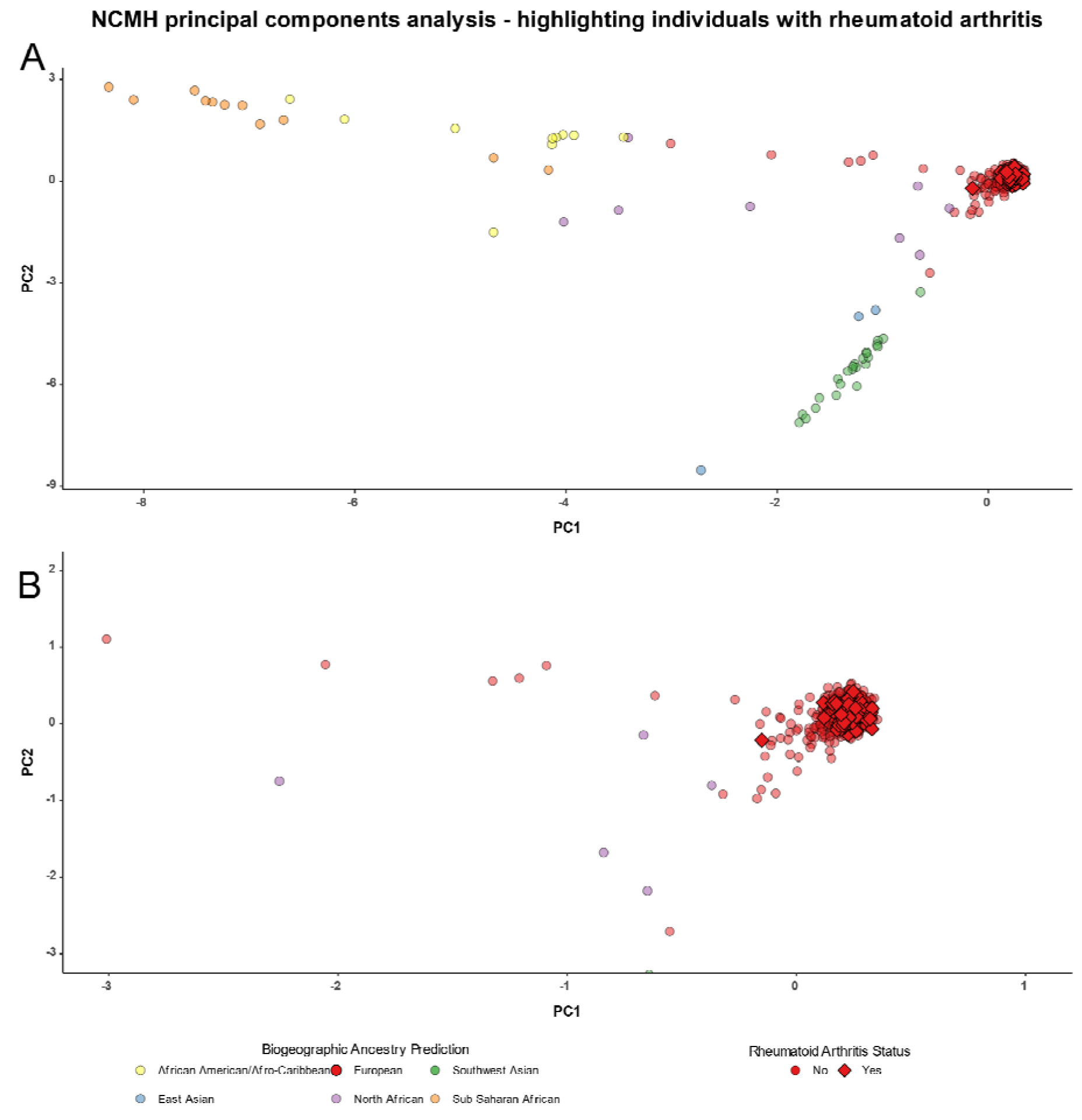
Principal components analysis of NCMH samples. A – All NCMH participants, with colours representing the individual’s biogeographical ancestry prediction derived from the DRAGON-Data QC pipeline. Diamonds represent individuals with SMI and a positive self-report of rheumatoid arthritis. B – represents a zoomed in version of A, focusing on the individuals with a positive history of rheumatoid arthritis. Only individuals shown in red were included in the analysis of any rheumatoid arthritis outcome in NCMH to avoid population stratification.

## Notes

### Competing Interest Statement

MJO, MCOD, and JTRW are supported by a collaborative research grant from Takeda Pharmaceuticals Ltd. for a project unrelated to work presented here. AFP, MJO, MCOD, and JTRW also reported receiving grants from Akrivia Health for a project unrelated to this submission. Takeda and Akrivia Health played no part in the conception, design, implementation, or interpretation of this study.

## References

1. Firth J, Siddiqi N, Koyanagi A, et al. The Lancet Psychiatry Commission: a blueprint for protecting physical health in people with mental illness. Lancet Psychiatry 2019; 6(8): 675–712.

2. Walker ER, McGee RE, Druss BG. Mortality in mental disorders and global disease burden implications: a systematic review and meta-analysis. JAMA Psychiatry 2015; 72(4): 334–41.

3. Correll CU, Solmi M, Veronese N, et al. Prevalence, incidence and mortality from cardiovascular disease in patients with pooled and specific severe mental illness: a large-scale meta-analysis of 3,211,768 patients and 113,383,368 controls. World Psychiatry 2017; 16(2): 163–80.

4. Launders N, Kirsh L, Osborn DPJ, Hayes JF. The temporal relationship between severe mental illness diagnosis and chronic physical comorbidity: a UK primary care cohort study of disease burden over 10 years. Lancet Psychiatry 2022; 9(9): 725–35.

5. Rodevand L, Rahman Z, Hindley GFL, et al. Characterizing the Shared Genetic Underpinnings of Schizophrenia and Cardiovascular Disease Risk Factors. Am J Psychiatry 2023; 180(11): 815–26.

6. Rodevand L, Bahrami S, Frei O, et al. Polygenic overlap and shared genetic loci between loneliness, severe mental disorders, and cardiovascular disease risk factors suggest shared molecular mechanisms. Transl Psychiatry 2021; 11(1): 3.

7. Rodevand L, Bahrami S, Frei O, et al. Extensive bidirectional genetic overlap between bipolar disorder and cardiovascular disease phenotypes. Transl Psychiatry 2021; 11(1): 407.

8. Bahrami S, Steen NE, Shadrin A, et al. Shared Genetic Loci Between Body Mass Index and Major Psychiatric Disorders: A Genome-wide Association Study. JAMA Psychiatry 2020; 77(5): 503–12.

9. Kendall KM, John A, Lee SC, et al. Impact of schizophrenia genetic liability on the association between schizophrenia and physical illness: data-linkage study. BJPsych Open 2020; 6(6): 1–7.

10. Arruda AL, Khandaker GM, Morris AP, Smith GD, Huckins LM, Zeggini E. Genomic insights into the comorbidity between type 2 diabetes and schizophrenia. Schizophrenia (Heidelb) 2024; 10(1): 22.

11. Reponen EJ, Ueland T, Rokicki J, et al. Polygenic risk for schizophrenia and bipolar disorder in relation to cardiovascular biomarkers. Eur Arch Psychiatry Clin Neurosci 2024; 274(5): 1223–30.

12. Veeneman RR, Vermeulen JM, Bialas M, et al. Mental illness and cardiovascular health: observational and polygenic score analyses in a population-based cohort study. Psychol Med 2024; 54(5): 931–9.

13. Bergstedt J, Pasman JA, Ma Z, et al. Distinct biological signature and modifiable risk factors underlie the comorbidity between major depressive disorder and cardiovascular disease. Nat Cardiovasc Res 2024; 3(6): 754–69.

14. Meijsen J, Hu K, Krebs MD, et al. Quantifying the relative importance of genetics and environment on the comorbidity between mental and cardiometabolic disorders using 17 million Scandinavians. Nat Commun 2024; 15(1): 5064.

15. Phulka JS, Ashraf M, Bajwa BK, Pare G, Laksman Z. Current State and Future of Polygenic Risk Scores in Cardiometabolic Disease: A Scoping Review. Circ Genom Precis Med 2023; 16(3): 286–313.

16. Segura AG, Martinez-Pinteno A, Gasso P, et al. Metabolic polygenic risk scores effect on antipsychotic-induced metabolic dysregulation: A longitudinal study in a first episode psychosis cohort. Schizophr Res 2022; 244: 101–10.

17. Alver M, Kasela S, Haring L, et al. Genetic predisposition and antipsychotic treatment effect on metabolic syndrome in schizophrenia: a ten-year follow-up study using the Estonian Biobank. Lancet Reg Health Eur 2024; 41: 100914.

18. Legge SE, Dennison CA, Pardiñas AF, et al. Clinical indicators of treatment-resistant psychosis. British Journal of Psychiatry 2020; 216(5): 259–66.

19. Lynham AJ, Hubbard L, Tansey KE, et al. Examining cognition across the bipolar/schizophrenia diagnostic spectrum. Journal of Psychiatry and Neuroscience 2018; 43(4): 245–53.

20. Wing JK, Babor T, Brugha T, et al. SCAN. Schedules for Clinical Assessment in Neuropsychiatry. Arch Gen Psychiatry 1990; 47(6): 589–93.

21. Apa APA. Diagnostic and Statistical Manual of Mental Disorders 4th edition: DSM-IV. Washington ed; 1994.

22. World Health O. ICD-10 : international statistical classification of diseases and related health problems : tenth revision. 2nd ed. Geneva: World Health Organization; 2004.

23. Underwood JFG, Kendall KM, Berrett J, et al. Autism spectrum disorder diagnosis in adults: Phenotype and genotype findings from a clinically derived cohort. British Journal of Psychiatry 2019; 215(5): 647–53.

24. Woolway GE, Legge SE, Lynham AJ, et al. Assessing the validity of a self-reported clinical diagnosis of schizophrenia. Schizophrenia (Heidelb) 2024; 10(1): 99.

25. Kuan V, Denaxas S, Gonzalez-Izquierdo A, et al. A chronological map of 308 physical and mental health conditions from 4 million individuals in the English National Health Service. Lancet Digit Health 2019; 1(2): e63–e77.

26. Lynham AJ, Knott S, Underwood JFG, et al. DRAGON-Data: a platform and protocol for integrating genomic and phenotypic data across large psychiatric cohorts. BJPsych Open 2023; 9(2): e32.

27. McCarthy S, Das S, Kretzschmar W, et al. A reference panel of 64,976 haplotypes for genotype imputation. Nat Genet 2016; 48(10): 1279–83.

28. Legge SE, Pardiñas AF, Helthuis M, et al. A genome-wide association study in individuals of African ancestry reveals the importance of the Duffy-null genotype in the assessment of clozapine-related neutropenia. Molecular psychiatry 2019; 24(3): 328–37.

29. Aragam KG, Jiang T, Goel A, et al. Discovery and systematic characterization of risk variants and genes for coronary artery disease in over a million participants. Nat Genet 2022; 54(12): 1803–15.

30. Mahajan A, Spracklen CN, Zhang W, et al. Multi-ancestry genetic study of type 2 diabetes highlights the power of diverse populations for discovery and translation. Nat Genet 2022; 54(5): 560–72.

31. Tsuo K, Zhou W, Wang Y, et al. Multi-ancestry meta-analysis of asthma identifies novel associations and highlights the value of increased power and diversity. Cell Genom 2022; 2(12): 100212.

32. Saevarsdottir S, Stefansdottir L, Sulem P, et al. Multiomics analysis of rheumatoid arthritis yields sequence variants that have large effects on risk of the seropositive subset. Ann Rheum Dis 2022; 81(8): 1085–95.

33. Evangelou E, Warren HR, Mosen-Ansorena D, et al. Genetic analysis of over 1 million people identifies 535 new loci associated with blood pressure traits. Nat Genet 2018; 50(10): 1412–25.

34. Graham SE, Clarke SL, Wu KH, et al. The power of genetic diversity in genome-wide association studies of lipids. Nature 2021; 600(7890): 675-9.

35. Demontis D, Walters RK, Martin J, et al. Discovery of the first genome-wide significant risk loci for attention deficit/hyperactivity disorder. Nature Genetics 2019; 51(1): 63–75.

36. Mullins N, Forstner AJ, O’Connell KS, et al. Genome-wide association study of more than 40,000 bipolar disorder cases provides new insights into the underlying biology. Nature Genetics 2021; 53(6): 817–29.

37. Howard DM, Adams MJ, Clarke T-K, et al. Genome-wide meta-analysis of depression identifies 102 independent variants and highlights the importance of the prefrontal brain regions. Nature Neuroscience 2019; 22(3): 343–52.

38. Trubetskoy V, Pardiñas AF, Qi T, et al. Mapping genomic loci implicates genes and synaptic biology in schizophrenia. Nature 2022; 604(7906): 502-8.

39. Murphy AE, Schilder BM, Skene NG. MungeSumstats: a Bioconductor package for the standardization and quality control of many GWAS summary statistics. Bioinformatics 2021; 37(23): 4593–6.

40. Wang Y, Namba S, Lopera E, et al. Global Biobank analyses provide lessons for developing polygenic risk scores across diverse cohorts. Cell Genom 2023; 3(1): 100241.

41. Ge T, Chen C-Y, Ni Y, Feng Y-CA, Smoller JW. Polygenic prediction via Bayesian regression and continuous shrinkage priors. Nature Communications 2019; 10(1): 1776.

42. Genomes Project C, Auton A, Brooks LD, et al. A global reference for human genetic variation. Nature 2015; 526(7571): 68-74.

43. Monti R, Eick L, Hudjashov G, et al. Evaluation of polygenic scoring methods in five biobanks shows larger variation between biobanks than methods and finds benefits of ensemble learning. Am J Hum Genet 2024; 111(7): 1431–47.

44. Martin Maechler PR, Christophe Croux, Valentin Todorov, Andreas Ruckstuhl,Matias Salibian-Barrera, Tobias Verbeke, Manuel Koller and Eduardo L. T. Conceicao, Maria Anna di Palma. robustbase: Basic Robust Statistics. R package version 0.99–3 ed; 2024.

45. Robinson CD, Tomek S, Schumacker R. Tests of Moderation Effects: Difference in Simple Slopes versus the Interaction Term. 2013; 2013.

46. Pillinger T, McCutcheon RA, Vano L, et al. Comparative effects of 18 antipsychotics on metabolic function in patients with schizophrenia, predictors of metabolic dysregulation, and association with psychopathology: a systematic review and network meta-analysis. Lancet Psychiatry 2020; 7(1): 64–77.

47. Allison P. Convergence Problems in Logistic Regression. Numerical Issues in Statistical Computing for the Social Scientist; 2003: 238–52.

48. van den Oord EJCG. Controlling false discoveries in genetic studies. Am J Med Genet B 2008; 147b(5): 637-44.

49. Viechtbauer W. Conducting Meta-Analyses in R with the metafor Package. Journal of Statistical Software 2010; 36(3): 1–48.

50. MacGregor-Fors I, Payton ME. Contrasting diversity values: statistical inferences based on overlapping confidence intervals. PLoS One 2013; 8(2): e56794.

51. Rees E, Owen MJ. Translating insights from neuropsychiatric genetics and genomics for precision psychiatry. Genome Medicine 2020; 12(1): 1–16.

52. Pizzol D, Trott M, Butler L, et al. Relationship between severe mental illness and physical multimorbidity: a meta-analysis and call for action. BMJ Ment Health 2023; 26(1).

53. Correll CU, Detraux J, De Lepeleire J, De Hert M. Effects of antipsychotics, antidepressants and mood stabilizers on risk for physical diseases in people with schizophrenia, depression and bipolar disorder. World Psychiatry 2015; 14(2): 119–36.

54. NHS-England. Implementing the early intervention in psychosis access and waiting time standard. 2023.

55. Solmi M, Fiedorowicz J, Poddighe L, et al. Disparities in Screening and Treatment of Cardiovascular Diseases in Patients With Mental Disorders Across the World: Systematic Review and Meta-Analysis of 47 Observational Studies. Am J Psychiatry 2021; 178(9): 793–803.

56. Chung R, Xu Z, Arnold M, et al. Using Polygenic Risk Scores for Prioritizing Individuals at Greatest Need of a Cardiovascular Disease Risk Assessment. J Am Heart Assoc 2023; 12(15): e029296.

57. Mbuya-Bienge C, Pashayan N, Kazemali CD, Lapointe J, Simard J, Nabi H. A Systematic Review and Critical Assessment of Breast Cancer Risk Prediction Tools Incorporating a Polygenic Risk Score for the General Population. Cancers (Basel) 2023; 15(22).

58. O’Sullivan JW, Ashley EA, Elliott PM. Polygenic risk scores for the prediction of cardiometabolic disease. Eur Heart J 2023; 44(2): 89–99.

59. Polcwiartek C, O’Gallagher K, Friedman DJ, et al. Severe mental illness: cardiovascular risk assessment and management. Eur Heart J 2024; 45(12): 987–97.

60. Osborn DP, Hardoon S, Omar RZ, et al. Cardiovascular risk prediction models for people with severe mental illness: results from the prediction and management of cardiovascular risk in people with severe mental illnesses (PRIMROSE) research program. JAMA Psychiatry 2015; 72(2): 143–51.

61. Murray GK, Lin T, Austin J, McGrath JJ, Hickie IB, Wray NR. Could Polygenic Risk Scores Be Useful in Psychiatry? JAMA Psychiatry 2021; 78(2): 210-.

62. Torkamani A, Wineinger NE, Topol EJ. The personal and clinical utility of polygenic risk scores. Nature Reviews Genetics: Nature Publishing Group; 2018. p. 581-90.

